# Microscopic dynamics modeling unravels the role of asymptomatic virus carriers in SARS-CoV-2 epidemics at the interplay between biological and social factors

**DOI:** 10.1101/2021.02.01.21250926

**Authors:** Bosiljka Tadić, Roderick Melnik

## Abstract

The recent experience of SARS-CoV-2 epidemics spreading revealed the importance of passive forms of infection transmissions. Apart from the virus survival outside the host, the latent infection transmissions caused by asymptomatic and presymptomatic hosts represent major challenges for controlling the epidemics. In this regard, social mixing and various biological factors play their subtle, but often critical, role. For example, a life-threatening condition may result in the infection contracted from an asymptomatic virus carrier. Here, we use a new recently developed microscopic agent-based modelling framework to shed light on the role of asymptomatic hosts and to unravel the interplay between the biological and social factors of these nonlinear stochastic processes. The model accounts for each human actor’s susceptibility and the virus survival time, as well as traceability along the infection path. These properties enable an efficient dissection of the infection events caused by asymptomatic carriers from those which involve symptomatic hosts before they develop symptoms and become removed to a controlled environment. Consequently, we assess how their relative proportions in the overall infection curve vary with changing model parameters. Our results reveal that these proportions largely depend on biological factors in the process, specifically, the virus transmissibility and the critical threshold for developing symptoms, which can be affected by the virus pathogenicity. Meanwhile, social participation activity is crucial for the overall infection level, further modulated by the virus transmissibility.

## I. INTRODUCTION

Epidemics spreading of new coronavirus in population is a collective social phenomenon. It arises from individual actors’ behaviour that can get infected and spread the infectious agents via multiplexity of contacts. In this regards, the human participation activity conditioned by daily mobility patterns has been recognised as a primary driving force for the epidemics spreading^1–3^. Contrary to many social processes, the epidemic spreading has a vital biological component, which is prominent at the elementary interactions scale^4,5^. In this respect, the recent developments with the SARS-CoV-2 epidemics have revealed several new features that were not previously recognised in virus spreading, see recent update in^6^. These are sizeable effects of the passive modes of the infection transmission^8–12^, which can be related to the virus biology and strongly individual susceptibility of the human hosts to this particular virus^13–16^. Specifically, these latent infection transmissions rely on the virus long survival time outside the human host^14,17^, which enables an indirect transmission to a new host. On the other hand, a considerable amount of asymptomatic hosts can remain unidentified^18–20^.

Since the very beginning of the SARS-CoV-2 epidemics, it has been revealed that a wide spectrum of symptoms may occur, from very mild or none, on one side, to severe symptoms and life-threatening pneumonia requiring ICU treatment and possible fatal outcome^14^. Apart from potential virus mutations over time, e.g., changed transmissibility and pathogenicity^11,21,22^, the observed individual susceptibility of human actors to the virus may range from a specific genetic origin to diverse factors related to the individual’s health condition^13,23,24^. The symptoms develop over a short period (1 to 5 days). Meanwhile, the asymptomatic cases spontaneously recover within a period of one to two weeks, changing infectiousness over time^25^. Even though their viral load varies differently in time^25,26^, both symptomatic and asymptomatic infected hosts are the virus carries and can spread the infection, cf. Fig. 1. Specifically, the circulating viruses produced by asymptomatic hosts can infect a susceptible individual, who, depending on the susceptibility, may or may not develop symptoms. Similarly, the viruses produced by presymptomatic carriers can lead to asymptomatic and symptomatic cases. Moreover, some measurements suggest that the viral contents are proportional to the severity of symptoms, and can vary over the infectious time^26^. Early estimates were that the proportion of asymptomatic cases could be as large as 80% of all infected^14^. A recent meta-analysis of the available data suggests a wide variation in the estimates from 20% up to 60% of asymptomatic carriers^20^.

**FIG. 1:**
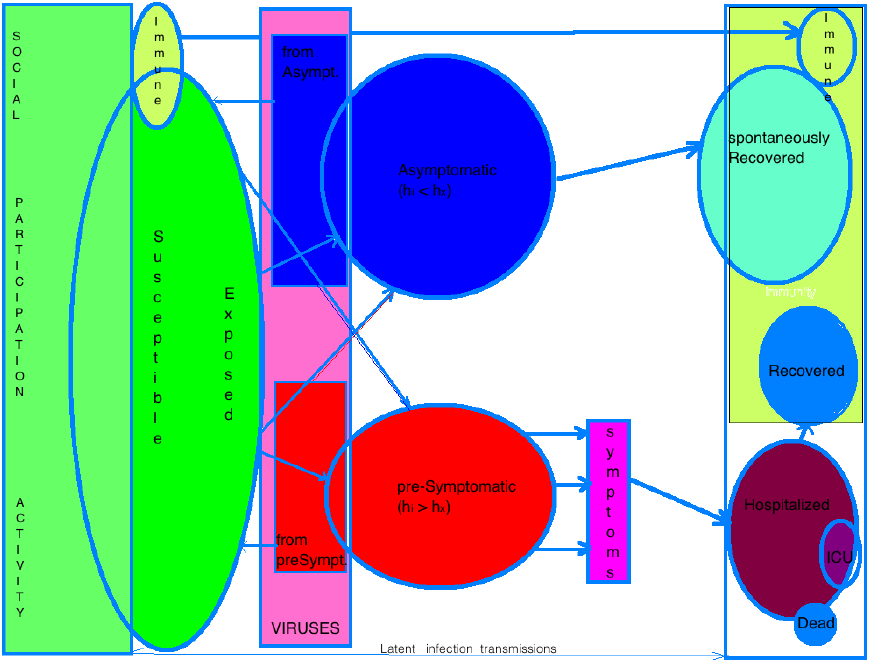
Schematic view of the processes: new susceptible individuals become exposed to active viruses. An infected individual is “asymptomatic”, if its susceptibility is low *h*_*i*_ < *h*_*x*_, or “presymptomatic” if its *h*_*i*_ ≥ *h*_*x*_, which means that the individual develops symptoms over time and subsequently becomes “hospitalised” and removed to a controlled environment. Whereas, “asymptomatic” carriers over time become “spontaneously recovered”. Meanwhile, the active circulating viruses consist of a group produced “by asymptomatic” and “by-presymptomatic.” hosts and can infect any one of the individuals, thus increasing either “asymptomatic” of “presymptomatic” group, depending on the agent’s susceptibility to the virus.

Given the occurrence of a sizable amount of asymptomatic virus carries, it is challenging to estimate the right parameters and predictions of the epidemics from the immediate data analysis^27,28^. Thus, it constitutes a considerable problem for efficient combat with the virus spreading^19,29,30^. The problem is increasingly more scientifically interesting in the third-wave epidemics because the virus circulates among a large fraction of the population. Hence, a better understanding of the factors that determine the proportion of the asymptomatic virus carries in a given social environment, and their impact on the overall infection growth is vital for managing the disease outbreak. In this work, we tackle these problems using the microscopic dynamics approach within the agent-based modelling framework developed in^1^.

Complementary to the standard mean-field models with continuous-time dynamics of interdependent equations for groups^31,32^, the microscopic agent-based modeling of SARS-CoV-2 epidemics gains an increasing attention^1,33–37^. This modelling approach provides us with intrinsic microscopic mechanisms of the epidemic processes, and the information on how it develops from the elementary interactions to the global-scale outcome. Moreover, it allows for considering individual features of the actors, their mobility and location of the interactions and participation in coupled stochastic processes^1,33,34,36**?**, 37^. We have recently developed an agent-based model^1^, where the human actors possess individual susceptibility to the virus. It thus allows us to differentiate between highly susceptible individuals, who can develop symptoms from those who are less susceptible and may become asymptomatically infected. The new host’s susceptibility accordingly modulates the probability of getting infected and the probability of that host to produce a new generation of viruses. Moreover, the model enables us to consider a finite survival time of the virus outside the host and different exposure times of each human actor. The process is visualised as a growing bipartite graphs, see Fig. 2, with host nodes who are producing the virus nodes (infected spots) during their infectious time and proportionally to the degree of infectiousness. A more detailed description of the model is given in the next section, see also^1^ for further details. This graphic representation of the process enables us to identify an infection path leading to each case. Therefore, in this modelling approach, each infection event where an actor encounters the virus is marked by the new host’s features and the host that produces the active virus.

**FIG. 2:**
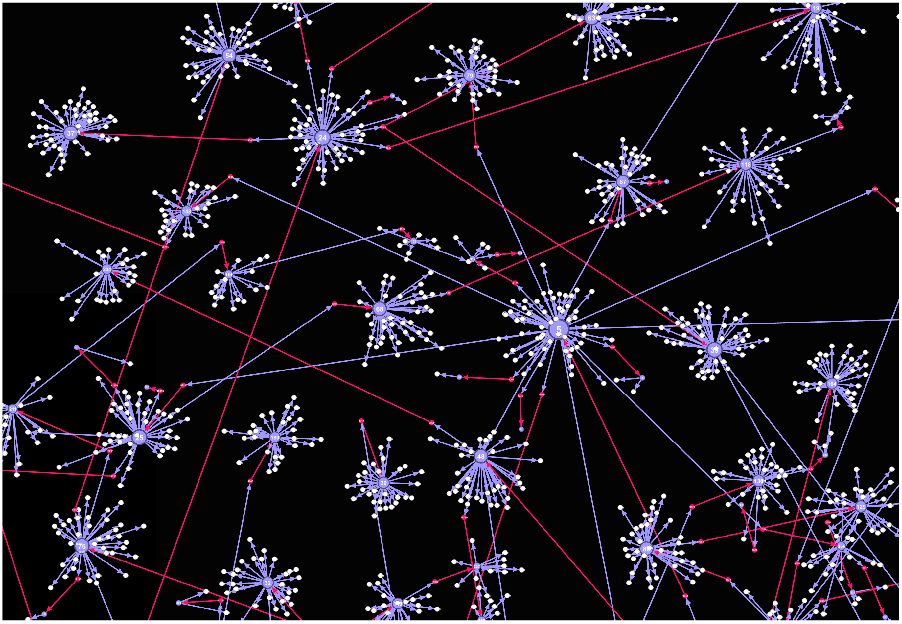
The close-up bipartite network of Host (large, blue) and Virus (small, white) nodes. Infection paths along the directed edges between successive Hosts occur via Virus nodes (shown in red).

In this work, we extend the developed model to keep the information about the preceding host’s susceptibility when the virus successfully hops to a new host. In this way, each infection case can be distinguished as either coming from a susceptible, i.e., potentially symptomatic case before it gets hospitalised or from a low-susceptibility host, which can be asymptomatic. The threshold susceptibility *h*_*x*_ is a varying parameter in our model, possibly assessable from some empirical data^13,18–20^. We note that these stochastic processes involving symptomatic and asymptomatic hosts are strongly entangled at the level of interactions, cf. Fig. 1, such that they can be suitably differentiated only at the microscopic scale with individual-based modelling. Precisely, by tracing every infection event, we can differentiate the virus original host and, thus, determine its contribution to the growth of the infectious curve over time. With the extensive simulations, we demonstrate how the asymptomatic host’s contribution to the infection growth varies with the social participation activity and the biological factors that determine the threshold susceptibility and transmission rate. Our simulations confirm that the overall infection level critically depends on social participation activity. Meanwhile, the biological factors are primarily responsible for the respective proportions of the asymptomatic and presymptomatic hosts in the overall infection curve.

## II. LATENT INFECTION TRANSMISSIONS: MODEL DETAILS

We adopt the model for latent infection transmissions developed in Ref.^1^. The process is visualised as an evolving bipartite graph consisting of infected individuals (Host nodes), who can produce viral spots (Virus nodes) during their infectious time, cf. Fig. 2. The model is driven by the *social participation* activity, which is approximated by an empirical time series *s*_*t*_ inferred from social networks^38^. In analogy to the time series of the human mobility within cities^35,39^, this time series represents a cumulative activity of an open social group in an area with circulating viruses. Thus, it does not assume any prior relationships among participants (see the Discussion section).

Features that influence the dynamics are individual characteristics of the human actors and viruses. Specifically, apart from a unique ID, *i*, each human actor has its individual *susceptibility* to the virus, *h*_*i*_ ∈ [0, 1] as well as a characteristic exposition to the viruses specified by its *exposure time* 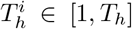. As mentioned in the Introduction, when a highly susceptible individual *h*_*i*_ ≥ *h*_*x*_ is infected it can develop symptoms over a certain number of days, 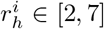; consequently, it gets hospitalised and moved to a controlled environment, and stops contributing to the latent infection transmission^1^. Meanwhile, individuals with the susceptibility below a threshold *h*_*i*_ *< h*_*x*_ would not develop symptoms by contracting the viruses; they will stay asymptomatically infected and eventually will spontaneously recover after *r*_*s*_ days. During the infectious time, both the asymptomatic and presymptomatic hosts produce viruses, presumably in different amounts^6,7^. In the model, the amount of viruses is proportional to the host’s susceptibility^1^. On the other hand, each virus node is characterised by a unique ID, survival time *T*_*v*_, and the information about the host that produced it. In^1^, by keeping track of the number of hops of the virus (virus generation) since the original infection case, its potential mutations can eventually moderate the transmission rate. For the present work, we keep the mutation factor fixed, *g* = 1. Other values can also be analysed with the developed framework, once reliable data on mutation patterns become available^22^. Each Virus node has a new property, *O*_*h*_, which is given by the susceptibility of the host that produced that virus.

As in the original model, in each event the basic transmission rate *λ*_0_ is modulated by the individual susceptibility of the agent *i* encountering the virus at the moment *t*, see Eq. (1). In addition, the probability 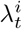 also varies in time due to the fluctuations in the global viral load *V* (*t*). Specifically,

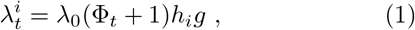

where the global feedback factor Φ_*t*_≡ *dV* (*t*)*/dH*_*a*_(*t*) follows the temporal fluctuations of the viral load with respect to the current number of active carriers *H*_*a*_(*t*). Note that the upper limit of the virus production rate at time *t* corresponds to a hypothetical situation where each active carrier has the maximum susceptibility *h*_*i*_ = 1 producing a new virus at every time step. Hence, the temporal feedback in the fluctuating transmission rate of Eq. (1) accounts for the actual heterogeneity of the virus carriers.

## III. MICROSCOPIC DYNAMICS AND SAMPLED QUANTITIES

As shown in Fig. 1, the social activity dynamics at an hourly resolution brings *s*_*t*_ new susceptible agents, which become exposed to active viruses. Note that the viruses can survive outside the hosts, such that the currently active viruses are those produced by all active carriers (asymptomatic as well as presymptomatic) within the past *T*_*v*_ = 4 hours. Each agent remains exposed for a period corresponding to its exposure time 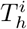, during which it can get infected with the probability given by Eq. (1). If infected, the agent is removed from the exposed agent’s list and appears in one of the infected agent’s groups, i.e., asymptomatic (if its susceptibility is below the threshold *h*_*x*_), or presymptomatic, if *h*_*i*_ ≥ *h*_*x*_. During their respective infectious times, the agents in both groups produce new viruses with a pace that is modulated by the agent’s susceptibility. After developing symptoms within an individual time interval of 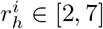 days, each symptomatic agent is hospitalised and removed from the process. Whereas, each asymptomatic agent stays in the process until its spontaneous recovery after *r*_*s*_ days (a parameter, equal to all agents). A detailed program flow is given as Supplementary information file in Ref.^1^.

As explained above, in the model, we keep information about the origin of each virus. Hence, in each new infection event, the number of infected *n*_*ta*_ increases by one if the virus originates from an asymptomatic host, and, in the case of a presymptomatic host, *n*_*ts*_ is increased. In each case the total number of infected agents per time step, *n*_*t*_ = *n*_*ta*_ + *n*_*ts*_ increases, however, the relative proportions of *n*_*ta*_ and *n*_*ts*_ can vary, depending on several parameters, as we show in the following. As it is schematically indicated in Fig. 1, these two processes are strongly interlinked at the microscopic scale. Particularly, the infection *by presymptomatic* hosts can end up as an asymptomatic as well as a presymptomatic case, depending on the susceptibility of the new arrival agent. Similarly, an infection *by asymptomatic* hosts can result in either an asymptomatic or symptomatic case; but each symptomatic case may eventually end up in the intensive care unit with an uncertain outcome, depending on its susceptibility^13,40^. Given the difficulty in detecting asymptomatic virus carriers in real life, understanding the intrinsic mechanisms along this potential line of events is of great importance.

This microscopic modelling framework allows us to keep full control of the process, which results in different time-varying quantities, as shown in Fig. 3. Specifically, for the time period spanning eight weeks and resolution of one hour, at each time step *i* = 1, 2 … *s*_*t*_ new agents are imported, and their individual properties *h*_*i*_ and 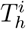 are fixed. Then at each time step, we determine the number of currently exposed agents *e*_*t*_, the number of agents infected from viruses by asymptomatic hosts, *n*_*ta*_, and by-presymptomatic hosts, *n*_*ts*_. By respecting the individual hospitalisation time for each presymptomatic host and spontaneous recovery time for all asymptomatic ones, as well as the virus survival time, we compute the number of active carriers *H*_*a*_(*t*) and active viruses *V* (*t*). Having these quantities at hand, we determine the actual transmission rate at each infection event, see Fig. 3.

**FIG. 3:**
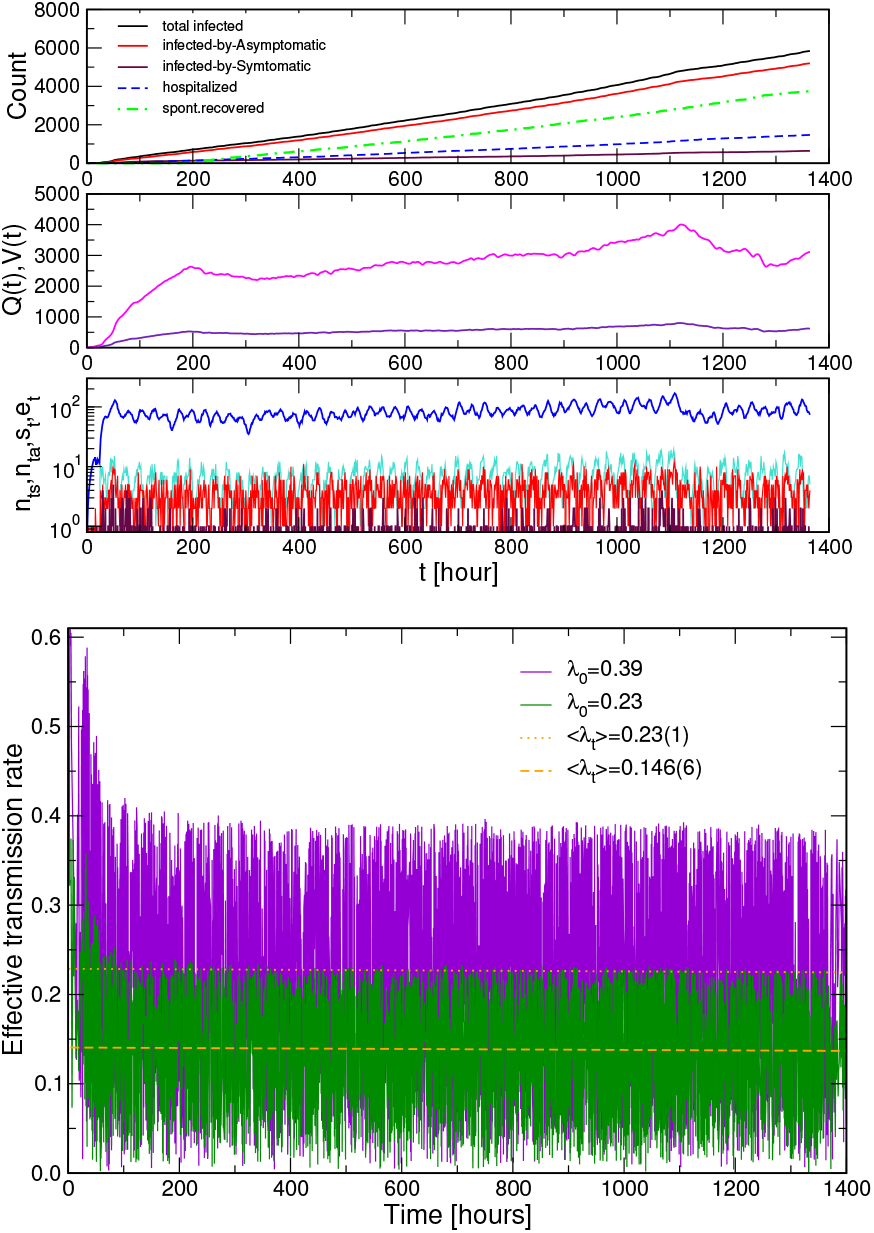
Temporal fluctuations of the social activity *s*_*t*_, the exposed agents *e*_*t*_, and the agents infected by asymptomatic *n*_*ta*_ and by presymptomatic *n*_*ts*_ hosts (bottom panel), and active carriers *H*_*a*_(*t*) and viruses *V* (*t*) (middle panel). The corresponding total number of infected *I*_*t*_ (infectious curve), and the proportions of infected by asymptomatic and by presymptomatic hosts, as well as the total number of hospitalised and spontaneously recovered over time, are shown in the top panel. In the simulations, we fixed *T*_*v*_ = 4 hours, maximum exposure time *T*_*h*_ = 24 hours, the spontaneous recovery time *r*_*s*_ = 7 days, and the maximum duration before developing symptoms *r*_*h*_ =7 days, the threshold value *h*_*x*_ = 0.8 and *λ*_0_ = 0.23. Bottom figure: The actual values of the transmission rate 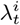 occurring in the sequence of infection events within the corresponding time frame, for two values of the basic rate *λ*_0_ indicated in the legend.

## IV. PROPORTION OF CASES INFECTED BY ASYMPTOMATIC CARRIERS FOR VARIED PARAMETERS

As shown above, the cases *infected by asymptomatic* hosts can be differentiated at the microscopic dynamics scale from the cases *infected by presymptomatic* hosts. Consequently, their relative contributions to the growth of the infectious curve can be systematically estimated. In the following, we focus on how these proportions vary in time (always starting from one symptomatic infected case), and how they depend on relevant parameters. Particularly, for a given social activity time series and fixed maximum exposure times of agents, we consider different values of the threshold susceptibility *h*_*x*_, the recovery time of asymptomatic hosts *r*_*s*_, and the basic transmission rate *λ*_0_. Specifically, for the same driving time series *s*_*t*_ as in Fig. 3, we show in Fig. 4 that the relative proportions of the cases infected by asymptomatic and by presymptomatic hosts strongly depend on the recognised threshold susceptibility. Meanwhile, the total number of infected remains statistically similar, being chiefly conditioned by the social participation activity. For example, for the threshold *h*_*x*_ = 0.8, corresponding to 80% asymptomatic cases, the fraction of infected by asymptomatic carriers levels up at 94% of all cases, whereas the remaining 6% cases are infected by presymptomatic virus carriers. The corresponding temporal evolution of these fractions is shown in the top-right panel of Fig. 5, starting from a single symptomatic case. On the other hand, by assuming that asymptomatic infections comprise of 40% of all cases, i.e., the threshold susceptibility *h*_*x*_ = 0.4, the total number of infected by presymptomatic carriers is initially higher, but the events attributed to infections by asymptomatic carriers win at later times. Eventually, for even lower values of the threshold, the number of infected by asymptomatic hosts remain below the number of infected by presymptomatic. The exact proportions evolve over time, as shown in Fig. 5, depending not only on the threshold value but also on the infectious time of the participants. The final outcomes (after eight weeks of evolution) are shown in the bottom panel of Fig. 4 for the whole range of threshold values and two infectious periods of the asymptomatic carriers, *r*_*s*_ = 7 and 14 days, respectively.

**FIG. 4:**
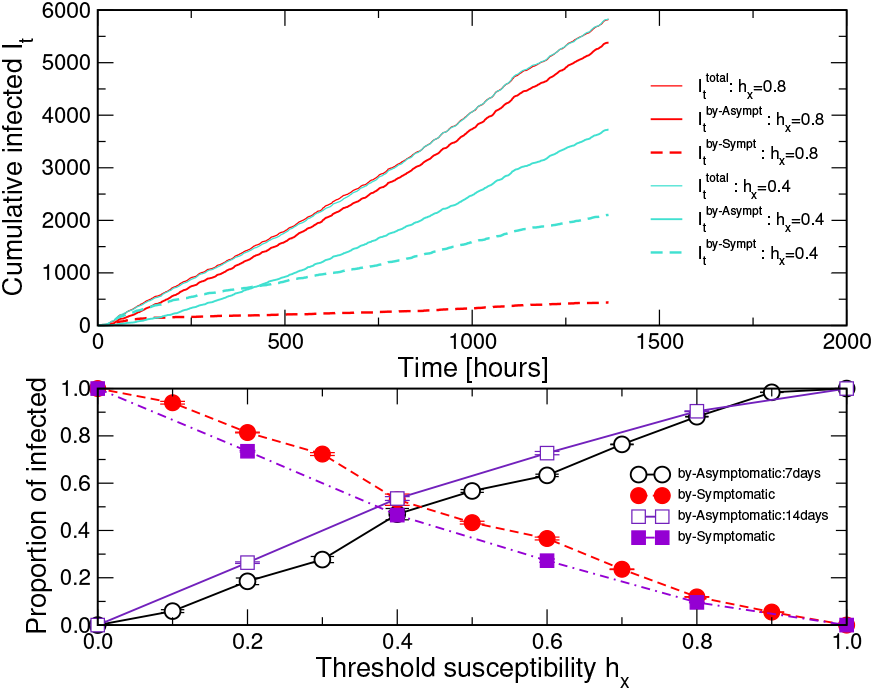
Top panel: Infection curve versus time (top blue line) and the respective proportions of the cumulative number of infected *by asymptomatic* hosts (red line) and *by presymptomatic* hosts (red dashed line) for the susceptibility threshold *h*_*x*_ = 0.8—corresponding to 80% of asymptomatic cases. The corresponding proportions (cyan full and dashed line, respectively) are for *h*_*x*_ = 0.4—corresponding to 40% of asymptomatic cases, meanwhile the total number of infected remains statistically the same. The recovery time of asymptomatic cases is 14 days, other parameters kept fixed (see text). Bottom panel: The proportions of infected by asymptomatic (open symbols) and by presymptomatic (filled symbols) plotted against the threshold susceptibility *h*_*x*_; two curves are for different recovery times of asymptomatic cases, 14 and 7 days, respectively.

**FIG. 5:**
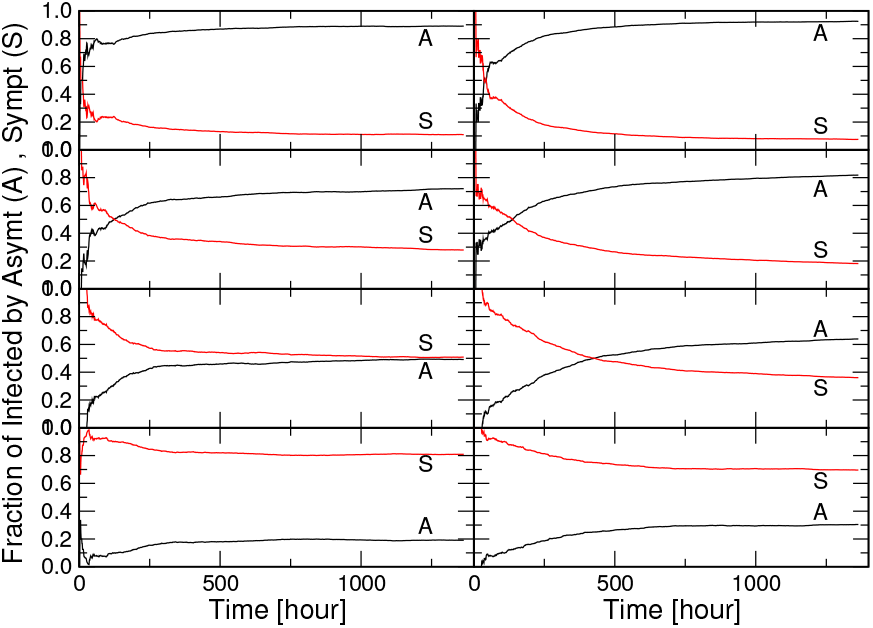
Time evolution of the proportions of the infections by the asymptomatic (black line) and presymptomatic (red line) cases for varied threshold susceptibility (top to bottom) *h*_*x*_ = 0.8,0.6, 0.4, and 0.2. Left column: corresponds to the situation with the spontaneous recovery time of asymptomatic cases 7 days, right column 14 days.

In the following, we examine how these proportions depend on the social participation level and the basic transmission rate. For this purpose, we extend the duration of the process. We use the same driving signal for the first eight weeks and then the signal with the reduced intensity but the same fractal structure for the following eight weeks, as shown in the top panel of Fig. 6. The simulation results for the infectious curves are shown in two bottom panels in Fig. 6. As this figure shows, the reduced social participation activity leads to the gradually slower growth of the total infection curve, in agreement with the findings in^1^ supporting the idea of social lockdown measures. Here, we are interested in how these variations in the social activity level combined with the transmission rate can affect the relative proportions of the infected by asymptomatic and by presymptomatic carriers. Specifically, we consider two cases of the threshold susceptibility, *h*_*x*_ = 0.8 and *h*_*x*_ = 0.2, and two basic transition rates, i.e., *λ*_0_ = 0.23 and, 70% increased transmission rate, *λ*_0_ = 0.39; the results are displayed in Fig. 6. These results reveal that, for a low basic transmission rate, even though the social activity level strongly influences the total number of cases, the *relative proportions* of these cases infected by asymptomatic and by presymptomatic carriers remain virtually unaffected. However, the increased basic transmission rate increases both the total number of infected and alters the proportions of the infected by asymptomatic and by presymptomatic carriers. Moreover, these proportions are dramatically different when the susceptibility threshold is high, e.g., 0.8, compared to the case when it is as low as 0.2. Particularly, in the first case, a practically entire increase of the infection curve for the increased transmission rate can be attributed to the infections by asymptomatic carriers. On the other hand, the situation is not symmetrical when the number of asymptomatic comprises 20% of all infected. In this case, we find that the proportions of infected by presymptomatic carriers exceed the proportion of infected by asymptomatic by an amount, which increases with the increased basic transmission rate.

**FIG. 6:**
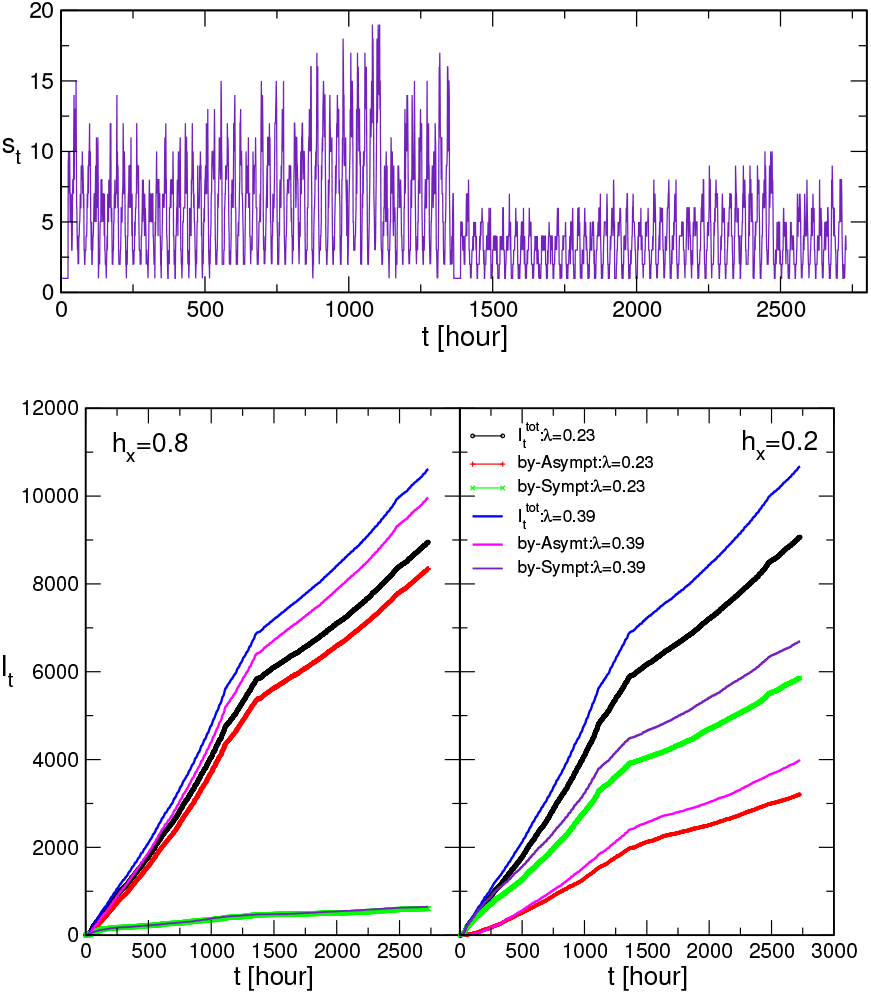
Top panel: Varied-intensity social participation activity time series *s*_*t*_. Bottom panels: For two threshold susceptibilities *h*_*x*_, shown in each panel, the total infection curve *I*_*t*_ and the corresponding proportions of infected by asymptomatic and by presymptomatic carriers for two different values of the basic transmission rate *λ*_0_=0.23 and 0.39.

## V. DISCUSSION AND CONCLUSIONS

We have studied the microscopic dynamics modelling of SARS-CoV-2 epidemics by building on the modelling framework developed in^1^. By keeping information about the host that produces a virus implicated in an infection event, we have been able to disentangle the cases attributed to asymptomatic from those caused by presymptomatic virus carriers. With the extensive simulations that comprise up to 16 weeks of the evolution time with the hourly resolution, we have demonstrated how the corresponding proportions of the infection curve vary in time and depend on the implicated bio-social factors. Dealing with a highly nonlinear stochastic process, we note that changing a parameter that affects the events at the microscopic scale may lead to an altered course of events and a different final outcome. At the same time, our results revealed certain regularities regarding the groups of social and biological factors. Specifically:

- The overall infection level critically depends on social participation activity. Hence, the increase of the infection curve can be forcefully controlled, e.g., by temporally reducing the social activity level, having the other factors fixed;
- For a given total infection, the relative ratio of the cases infected by asymptomatic carriers to the cases infected by presymptomatic carriers crucially depends on several biological factors. These are, the threshold susceptibility (depending on the virus pathogenicity and human genetic and other health factors of the implicated actors), and the virus transmissivity;
- The interplay between social and biological factors can be altered, increasing the proportion of cases attributed to the asymptomatic carriers, when the virus transmissivity considerably increases. For example, the considered situation where the basic transmission rate is increased by 70% is motivated by recently debated potential mutations of SARS-CoV-2, see^21,22^.

In conclusion, our microscopic dynamics modelling of the SARS-CoV-2 epidemics reveals the interplay between different biological and social factors of this nonlinear process, which shapes the increase of the infectious curve and the proportions attributed to asymptomatic and presymptomatic virus carriers. Given social participation activity under control, our results shed light on the intrinsic action mechanisms of the key biological factors. In particular, these are the critical threshold susceptibility to the virus and the increased virus transmissivity, which lead to the increased proportions of the infections by asymptomatic carriers. Thus, assuming that pertinent empirical data on the virus transmissivity and pathogenicity can be available for a particular population, these findings should assist in better estimates of the impact of the hidden asymptomatic carriers, and consequently in the design of the appropriately improved measures.

## Data Availability

This study is purely theoretical and does not include any experimental or clinical trials. Used information on SARS-CoV-2 epidemics is publicly available.

## Acknowledgments

B.T. acknowledges the financial support from the Slovenian Research Agency (research code funding number P1-0044). R.M. thanks to the NSERC and the CRC Program for their support and he is also acknowledging the support of the BERC 2018-2021 program and Spanish Ministry of Science, Innovation, and Universities through the Agencia Estatal de Investigacion (AEI) BCAM Severo Ochoa excellence accreditation SEV-2017-0718, and the Basque Government fund “AI in BCAM EXP. 2019/00432”

## References

1 B. Tadić and R. Melnik. Modeling latent infection transmissions through biosocial stochastic dynamics. PloS one, 15(10):e0241163–16, 2020.

2 F. Brauer. Mathematical epidemiology: past, present and future. Infectious Disease Modelling, 2:113–127, 2017.

3 M. Schneider, J. R. Johnson, N. J. Krogan, and S. K. Chanda. Chapter 12 - the virushost interactome: Knowing the players to understand the game. In M. G. Katze, M. J. Korth, G. L. Law, and N. Nathanson, editors, Viral Pathogenesis (Third Edition), pages 157 – 167. Academic Press, Boston, third edition, 2016.

4 R. W. Doms. Chapter 3 - basic concepts: A step-by-step guide to viral infection. In M. G. Katze, M. J. Korth, G. L. Law, and N. Nathanson, editors, Viral Pathogenesis (Third Edition), pages 29 – 40. Academic Press, Boston, third edition edition, 2016.

5 M. T. Ferris, M. T. Heise, and R. S. Baric. Chapter 13 - Host genetics: It is not just the virus, stupid. In M. G. Katze, M. J. Korth, G. L. Law, and N. Nathanson, editors, Viral Pathogenesis (Third Edition), pages 169 – 179. Academic Press, Boston, third edition edition, 2016.

6 M. Cevik, K. Kuppalli, J. Kindrachuk, and M. Peiris. Virology, transmissions, and pathogenesis of sars-cov-2. BMJ, 371, 2020.

7 J. Dufloo, L. Grzelak, I. Staropoli et al.. Asymptomatic and symptomatic SARS-CoV-2 infections elicit polyfunctional antibodies. medRxiv:https://www.medrxiv.org/content/10.1101/2020.11.12.20230508v1.full.pdf, 2020

8 N. Xiong, T. Wang, and Z. Lin. Invisible spread of sars-cov-2. The Lancet Infectious Diseases, 20(9):1011–1012, 2020.

9 C.E. Enyoh, A.W. Verla, W. Qingyue, D.K. Yadav, M.A. Hossain Chowdhury, B.O. Isiuku, T. Chowdhury, F.C. Ibe, E.N. Verla, and T.O. Maduka. Indirect exposure to novel coronavirus (sars-cov-2): An overview of current knowledge. Preprints 2020, art.2020040460, 2020.

10 J. Cai, W. Sun, J. Huang, M. Gamber, J. Wu, and G. He. Indirect virus transmission in cluster of covid-19 cases, wenzhou, china, 2020. Emerging infectious diseases, 26(6):13431345, 2020.

11 J. Chen. Pathogenicity and transmissibility of 2019-nCoVa quick overview and comparison with other emerging viruses. Microbes and Infection, 22(2):69 –71, 2020.

12 M. Cevik, M. Tate, O. Lloyd, A. E. Maraolo, J. Schafers, and A. Ho. SARS-CoV-2, SARS-CoV, and MERS-CoV viral load dynamics, duration of viral shedding, and infectiousness: a systematic review and meta-analysis. The Lancet Microbe, 2(1):e13–e22, 2021.

13 F. Wang, S. Huang, R. Gao, Y. Zhou, C. Lai, Z. Li, et al. Initial whole-genome sequencing and analysis of the host genetic contribution to covid-19 severity and susceptibility. Cell Discovery, 6:83, 2020.

14 Y. Wang, Y. Wang, Y. Chen, and Q. Qin. Unique epidemiological and clinical features of the emerging 2019 novel coronavirus pneumonia (covid-19) implicate special control measures. Journal of Medical Virology, 92(6):568–576, 2020.

15 S. Nersisyan, N. Engibaryan, A. Gorbonos, K. Kirdey, A. Makhonin, and A. Tonevitsky. The potential role of miRNAs-21-3p in coronavirus-host interplay. PeerJ, 8:e994, 2020.

16 Y. Zhang, X. Geng, Y. Tan, Q. Li, C. Xu, J. Xu, et al. New understanding of the damage of sars-cov-2 infection outside the respiratory system. Biomedicine & Pharmacotherapy, 127:110195, 2020.

17 A.W.H. Chin, J.T.S. Chu, M.R.A. Perera, K.P.Y. Hui, H-L. Yen, M.C.W. Chan, M. Peiris, and L.L.M. Poon. Stability of sars-cov-2 in different environmental conditions. Lancet Microbe, 1(1):e10, 2020.

18 K. J. Meyers, M. E. Jones, I. A. Goetz, F. T. Botros, J. Knorr, D. H. Manner, and B. Woodward. A crosssectional community-based observational study of asymptomatic sars-cov-2 prevalence in the greater indianapolis area. Journal of medical virology, 92(11):28742879, 2020.

19 E. Meyerowitz, A. Richterman, I. Bogoch, N. Low, and M. Cevik. Towards an accurate and systematic characterization of persistently asymptomatic infection with sars-cov-2. Lancet Inf. Disease, ISSN 1473-3099, 2020.

20 D. Buitrago-Garcia, D. Egli-Gany, M. J. Counotte, S. Hossmann, H. Imeri, A. M. Ipekci, G. Salanti, and N. Low. Occurrence and transmission potential of asymptomatic and presymptomatic sars-cov-2 infections: A living systematic review and meta-analysis. PLOS Medicine, 17(9):1–25, 2020.

21 L. van Dorp, D. Richard, C.C.S. Tan, L.P. Shaw, M. Acman, and F. Ballox. No evidence for increased transmissibility from recurrent mutations in sars-cov-2. Nature Communications, 11:5986, 2020.

22 E. Callawey. Making sense of coronavirus mutations. Nature, 585:174–177, 2020.

23 C. Lu, R. Gam, A.P. Pandurangan, and J. Gough. Genetic risk factors for death with sars-cov-2 from the uk biobank. medRxiv art.2020.07.01.20144592, 2020.

24 Risk factors for death from covid-19. Nature Reviews Immunology, 20:407, 2020.

25 X. He, E.H.Y. Lau, and P. et al. Wu. Temporal dynamics in viral shedding and transmissibility of covid-19. Nature Medicine, 26:672–675, 2020.

26 K. K.-W. To, O. T.-Y. Tsang, W.-S. Leung, A. R. Tam, T.-C. Wu, D. C. Lung, et al. Temporal profiles of viral load in posterior oropharyngeal saliva samples and serum antibody responses during infection by sars-cov-2: an observational cohort study. The Lancet Infectious Diseases, 20(5):565 – 574, 2020.

27 C. Anastassopoulou, L. Russo, A. Tsakris, and C. Siettos. Data-based analysis, modelling and forecasting of the covid-19 outbreak. PLOS ONE, 15(3):1–21, 2020.

28 P. Magal and G. Webb. Predicting the number of reported and unreported cases for the covid-19 epidemic in South Korea, italy, France and Germany. medRxiv, 2020.03.21.20040154, 2020.

29 X. Qiu, A.I. Nergiz, A.E. Maraolo, I.I. Bogoch, N. Low, and M. Cevik. Defining the role of asymptomatic and presymptomatic sars-cov-2 transmission a living systematic review. medRxiv, 2020.09.01.20135194v2, 2020.

30 M. Yanes-Lane, N. Winters, F. Fregonese, M. Bastos, S. Perlman-Arrow, J. R. Campbell, and D. Menzies. Proportion of asymptomatic infection among covid-19 positive persons and their transmission potential: A systematic review and meta-analysis. PLOS ONE, 15(11):1–21, 2020.

31 G. Giordano, F. Blanchini, R. Bruno, P. Colaneri, A. Di Filippo, A. Di Matteo, and M. Colaneri. Modelling the covid-19 epidemic and implementation of populationwide interventions in italy. Nature Medicine, 26(6):855– 860, 2020.

32 A. J. Kucharski, T. W. Russell, C. Diamond, Y. Liu, J. Edmunds, S. Funk, et al. Early dynamics of transmission and control of covid-19: a mathematical modelling study. The Lancet Infectious Diseases, 20(5):553 – 558, 2020.

33 E. Cuevas. An agent-based model to evaluate the covid-19 transmission risks in facilities. Computers in Biology and Medicine, 121:103827, 2020.

34 C. Wolfram. An agent-based model of covid-19. Complex Systems, 29:87–105, 2020.

35 S. A. Müller, M. Balmer, W. Charlton, R. Ewert, A. Neumann, C. Rakow, T. Schlenther and K. Nagel. A realistic agent-based simulation model for COVID-19 based on a traffic simulation and mobile phone data. 2011.11453v1, 2020.

36 Z. Burda. Modelling excess mortality in covid-19-like epidemics. Entropy, 22(11), e1236, 2020.

37 S.L. Chang, N. Harding, C. Zacherson, O.M. Cliff, and M. Prokopenko. Modelling transmission and control of the covid-19 pandemic in australia. Nat. Communications, 11:5710, 2020.

38 M. Šuvakov, M. Mitrović, V. Gligorijević, and B. Tadić. How the online social networks are used: dialogues-based structure of myspace. Journal of the Royal Society Interface, 10(79):20120819, 2012.

39 M.C. Gonzales, Y. Xu, and A. Hernando. Unraveling the interplay of the urban form, mobility and social mixing in the light of the covid-19 pandemic. Book of abstracts, pp. 138, Conference on Complex Systems CCS2020-ONLINE, 2020.

40 C. Lu, R. Gam, A. P. Pandurangan, and J. Gough. Genetic risk factors for death with sars-cov-2 from the uk biobank. medRxiv, 2020.07.01.20144592, 2020.

